# Prolonged survival of a patient with active MDR-TB HIV co-morbidity: Insights from a Mycobacterium tuberculosis strain with a unique genomic deletion

**DOI:** 10.1101/2023.08.28.23294468

**Authors:** Mor Rubinstein, Andrei Makhon, Yelena Losev, Gal Zizelski Valenci, Yair E. Gatt, Hanah Margalit, Ephraim Fass, Ina Kutikov, Omer Murik, David A. Zeevi, Michal Savyon, Luba Tau, Hasia Kaidar Shwartz, Zeev Dveyrin, Efrat Rorman, Israel Nissan

## Abstract

Coinfection of HIV and multidrug-resistant tuberculosis (MDR-TB) presents significant challenges in terms of the treatment and prognosis of tuberculosis, leading to complexities in managing the disease and impacting the overall outcome for TB patients. This study presents a remarkable case of a patient with MDR-TB and HIV coinfection who survived for over eight years, despite poor treatment adherence and comorbidities. Whole genome sequencing (WGS) of the infecting *Mycobacterium tuberculosis* (*Mtb*) strain revealed a unique genomic deletion, spanning 18 genes, including key genes involved in hypoxia response, intracellular survival, immunodominant antigens and dormancy. This deletion, that we have called “Del-X”, potentially exerts a profound influence on the bacterial physiology and its virulence. Only few similar deletions were detected in other non-related *Mtb* genomes worldwide. *In vivo* evolution analysis identified drug resistance and metabolic adaptation mutations and their temporal dynamics during the patient’s treatment course.

## Introduction

Tuberculosis (TB) is a major cause of morbidity and one of the leading causes of death worldwide [1]. TB/HIV coinfection and drug resistant TB pose challenges to the TB elimination efforts. Effective treatment of TB, particularly multidrug-resistant TB (MDR-TB) and extensively drug resistant TB (XDR-TB) is long, expensive and toxic. The long treatment duration (6-22 months [2]), and severe adverse effect of anti MDR-TB medications often lead to poor patient adherence to the treatment, putting not only these patients, but also the people in their close proximity, at risk [3,4]. Discontinuous treatment and partial treatment are two leading causes for drug resistance [5].

Coinfection of TB and HIV exacerbates the symptoms of both diseases in a synergetic manner, as both pathologies impair the immune system’s ability to defend against these pathogens [6]. Coinfection with HIV significantly increases mortality among TB patients [7]. For example, a study conducted in Eastern Europe reported a median survival time of 5.9 years following diagnosis with MDR-TB or XDR-TB, which decreases to 1.9 years for patients also diagnosed with HIV [8]. Additional risk factors for death among TB patients include alcoholism, male gender and old age, among others [7,9].

*Mycobacterium tuberculosis (Mtb)*, the major causative agent of TB, can exist inside the human host either in an active, replicating state, or in a dormant state. Dormancy is a programed response which enables *Mtb* to survive inside macrophage phagosomes under conditions of starvation, hypoxia, low pH, and oxidative and nitrosative stress. During dormancy, bacteria cease replication, down-regulate central metabolism and switch to anaerobic metabolism [10]. The induction of dormancy is regulated by the dormancy survival DosR regulon [11,12]. This dormant state allows latent infection that can persist for many years or even a lifetime without clinical symptoms. However, if a critical number of bacteria manage to evade the host immune system and resume an active, replicating state, the patient develops an active, symptomatic TB disease [13,14].

The chronic nature of infection with *Mtb* allows for within-host evolution of the infecting bacteria over time. This process involves the accumulation of mutations, which are believed to contribute to the pathogen’s adaptation to the environmental conditions within the host. Indirect evidence supporting the selective advantage of these mutations is the observation that they often occur in the same genes across different patients, suggesting homoplastic evolution. A major mycobacterial function influenced by these mutations is drug resistance, frequently accompanied by compensatory mutations that mitigate the fitness cost of drug resistance mutations. Genes linked to virulence, lipid and cell wall metabolism and glycerol-3-phosphate metabolism have also been demonstrated to accumulate mutations during host infection [15,16].

Compared to other bacteria, the genome of *Mtb* is highly conserved. Modern *Mtb* strains do not typically acquire new genomic features through horizontal gene transfer [17,18]. However, *Mtb* strains often lose fragments of their genomes. Baena *et al* found 4111 genomic deletions longer than 1000 bp in 522 *Mtb* isolates of L4 lineage [19]. One of those deletions, that is 6479 bp long and results in the loss of 10 genes, was observed in 249 genomes from four different countries. Other large deletions were also detected in multiple strains. Most of these strains were obtained from sputum, which is produced exclusively during active respiratory tuberculosis. Assuming each unique deletion originated from a distinct event, it is therefore evident that some large genomic deletions do not impede pathogeny and transmission.

Israel has maintained ongoing surveillance of all TB cases since 1997. Patients with suspected pulmonary TB in Israel are requested to provide sputum or other biological specimens for culture. All isolates are sent by primary laboratories to the National Mycobacterium Reference Laboratory (NMRL) in Tel Aviv, Israel. NMRL identifies the strain, performs drug susceptibility tests and maintains a bank of strains as part of routine work. Since 2019, all new TB strains are fully sequenced (WGS) for epidemiological purposes and for drug susceptibility prediction. Between 2010 and 2019, NMRL received 35 TB samples from a patient diagnosed with both TB and HIV. These samples were found to belong to a single clone of MDR-TB characterized by a unique genomic deletion spanning 18 genes. In spite of the patient’s long progressive AIDS and active TB, incomplete treatment, and multiple risk factors, this patient survived for almost 9 years. Here we describe this exceptional case, and discuss the distinct features of the infecting pathogen that may have contributed to this survival.

## Results

### Case description

A man in his late 50’s was diagnosed with multidrug resistant tuberculosis (MDR-TB) in 2010, five years after being diagnosed with HIV. Over the next eight and a half years, he suffered from this severe and notorious co-morbidity, and eventually passed away in 2019 at his late 60’s (Figure 1). The recorded causes of deaths included TB, alcoholism, AIDS and acute respiratory failure.

**Figure 1:**
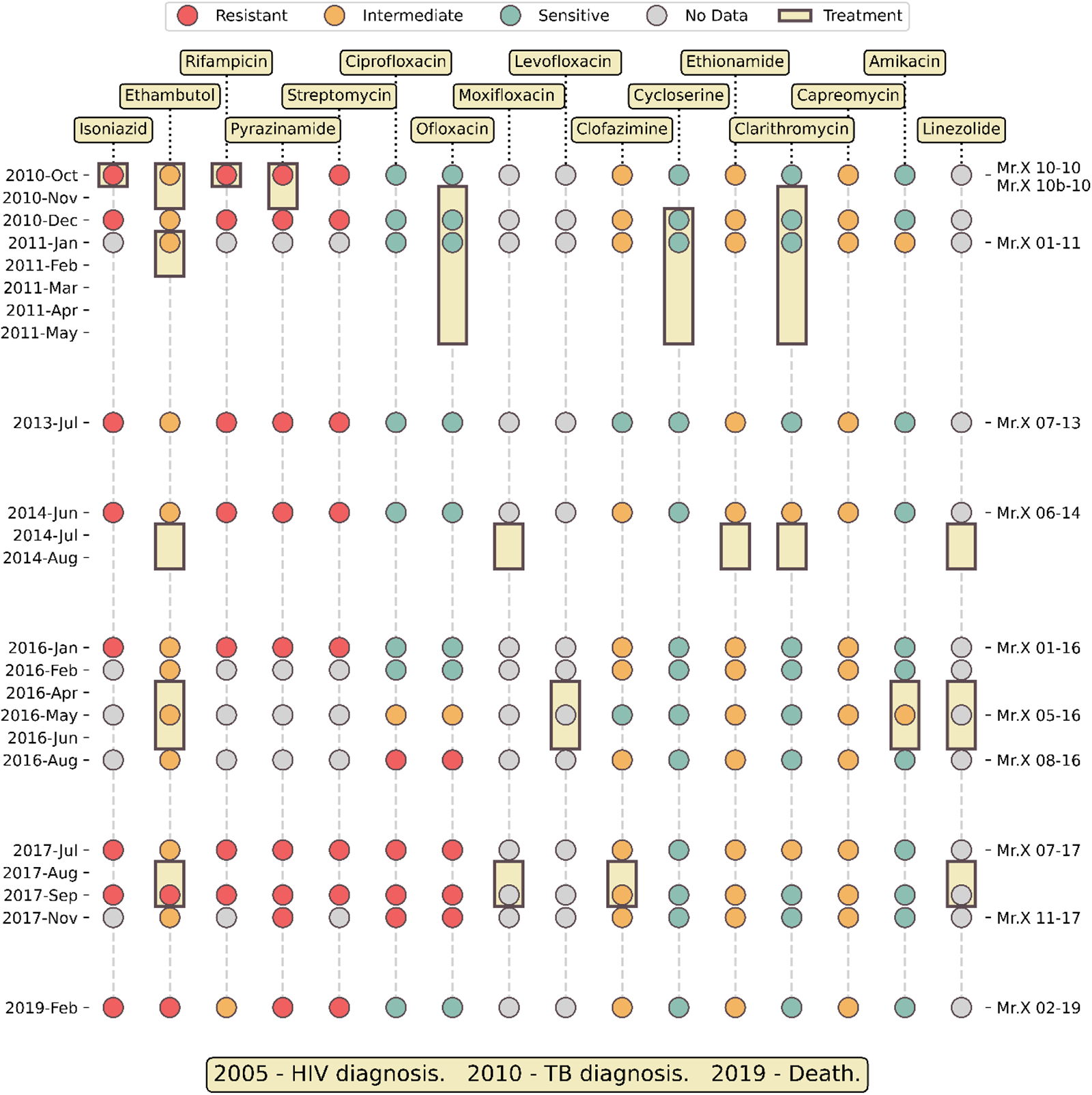
Timeline medical history of Mr. X. Colored dots represent susceptibility to antibiotics of the isolates. Yellow rectangles show the duration of antimicrobial treatment with a specific antibiotic. The ticks on the right side indicate the sequenced samples.

The patient, who will be referred to as ‘Mr. X’ from this point on, was born in Russia and immigrated to Israel. Shortly after his immigration he was diagnosed with tuberculosis. His compliance with medical treatment was poor and sporadic, therefore his medical records were sparse, difficult to track and incomplete.

Throughout the years, Mr. X started treatment multiple times, either in community settings or during hospitalization. Importantly, he never completed any of his therapies and often disappeared from the medical authorities’ radar. The 35 TB positive sputum samples collected in each of his therapy were sent to NMRL and tested for anti-TB drugs susceptibility (Figure 1, Supplementary Table S1). Eleven samples were subjected to Whole Genome Sequencing (WGS), which confirmed that the infection comprised a single clone of *Mtb* Beijing lineage throughout the entire illness duration.

Initially, Mr. X was treated using standard therapy for sensitive TB, i.e. a combination of rifampicin (RIF), isoniazid (INH), ethambutol (EMB) and pyrazinamide (PZA) (Figure 1). However, after the laboratory reported that the infecting strain is resistant to RIF, INH and PZA, these drugs were replaced, and within two months of the first TB diagnosis, his therapy regimens included primarily second line drugs. Overall, he was treated with 13 different anti-tuberculosis antibiotics (Figure 1).

Despite his HIV infection, Mr. X firmly refused antiretroviral therapy (ART). In 2017, his CD4 count was around 80/microliter, corresponding to 10% of lymphocytes, with CD4/CD8 ratio lower than 0.2 and a viral load near 350,000 per ml. In the following year his tests revealed a CD4 count around 80 per microliter (10%), CD4/CD8 ratio lower than 0.2 and viral load around 900,000 per ml. These tests indicate an active AIDS, with a severely compromised immune system.

In light of his HIV and MDR-TB comorbidity, his poor compliance to treatment, his age, male gender and alcoholism, the survival of this patient around nine years is very unusual.

### Drug resistance mutations

Drug Susceptibility Testing (DST) results are shown in Figure 1 and Supplementary Table S1. Antibiotic resistance was also predicted based on whole genome sequencing (WGS), by mapping short reads of each TB isolate to the reference genome, and comparing the variations identified to a database of known resistance mutations [20]. The genomic resistance prediction generally agreed with the phenotypic resistance determined by DST. We identified mutations that are known to confer resistance to INH, RIF, EMB and STM (Supplementary Table S2). The situation was different for PZA resistance. The gene *pncA* encodes an enzyme required for the transformation of PZA into its active form, pyrazinoic acid. Genetic resistance to PZA is usually caused by mutations in this gene [20]. However, while DST results showed PZA resistance, no mutation was found that could account for it. Upon careful inspection, we noticed there was no coverage of the genomic region around *pncA* by the strain’s short reads. This uncovered region proved to be a genomic deletion of size 15.7 kb (Del-X, see next section, Figure 2). The lack of functional *pncA* gene accounted for the PZA resistance observed by DST. The acquisition of an A90V mutation in *gyrA*, explains the transition to fluoroquinolones (FQs) resistance, shown in Figure 1. This mutation, which is known to confer FQ resistance, was not present in any of the isolates until May 2016. It first appeared in August 2016 and was present in the 2017 isolates, but disappeared in the genome of the last, FQs sensitive, sample taken in 2019. A frameshift mutation in the gene *ethA*, classified by the WHO as associated with ‘ethionamide resistance – interim’, was found in the genomes of 8 out of the 11 sequenced samples. A careful inspection revealed a significant coverage of this mutation in some reads of an additional sequenced sample. The significance of *ethA* to ethionamide (ETO) resistance is similar to that of *pncA* to PZA resistance: it alters this pro-drug to its active form. Nevertheless, this mutation only provided intermediate resistance (Figure 1). Two of the sequenced genomes did not have this mutation, but had another *ethA* mutation of uncertain significance, which was not shared by the other samples (Supplementary Table S2). In contrast to genomic prediction, no differences in ETO DST were detected among the different samples. No clofazimine (CFZ) resistance mutation was identified to explain the intermediate resistance observed in most isolates. We identified one mutation with uncertain significance regarding capreomycin (CAP) and amikacin (AMK) resistance in all samples. We were unable to explain the differences among AMK DST results using genomic data. Genomic-based susceptibility states were not predicted for cycloserine (CYC) and clarithromycin (CLR).

**Figure 2:**
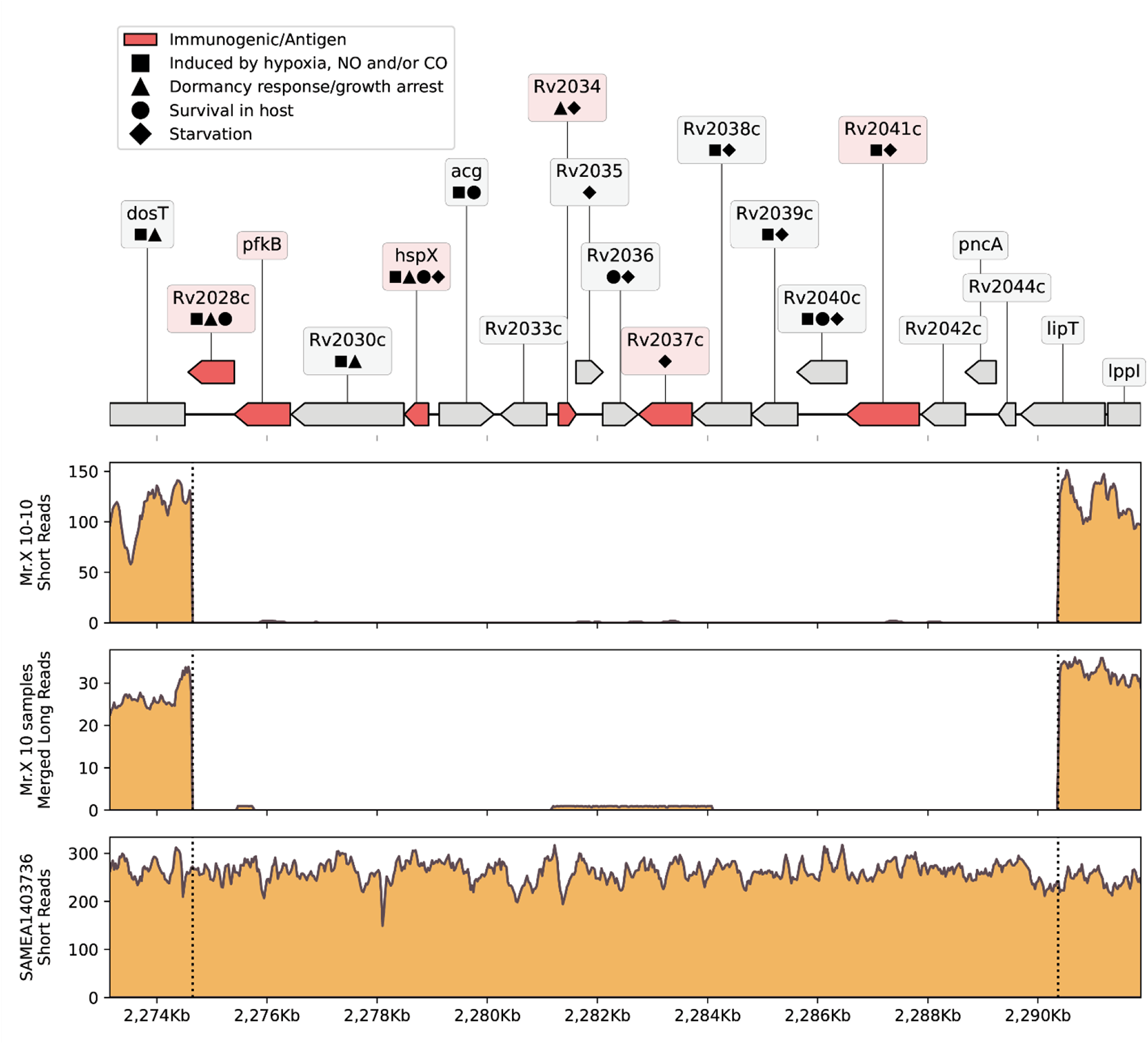
Del-X genes and coverage. On the top: a genomic map of the Del-X region according to the *Mtb* H37Rv reference genome. Genes in red are immunogenic. Symbols represent gene functions of interest. On the bottom: coverage of the Del-X region with short Illumina reads of sample “Mr.X 10-10”; combined coverage of all Nanopore long reads from Mr. X’s samples except for “Mr.X 02-19”; coverage of the isolate that is most closely related to Mr. X’s samples, according to Pathogen Detection. Dashed lines represent deletion borders, and left labels on ticks show coverage depth. BWA-MEM [72] was used to map short reads, and minimap2 v2.24-r1122 [85] was used to map merged long reads.

### Del-X

As mentioned above, WGS of the *Mtb* strain from Mr. X revealed a genomic deletion of size 15.7 kb, spanning bases 2274648 to 2290386 when compared to the reference *Mycobacterium tuberculosis* strain H37Rv (RefSeq id NC_000962.3, Figure 2). We have designated this deletion as Del-X. The precise boundaries of Del-X were verified using both short and long read sequencing technologies (Figure 2). The reference genome contains 18 genes in this region. These genes play important roles in the bacterial response to hypoxia, antibiotics susceptibility, interaction with the host, intracellular survival and dormancy. Many of them are considered immunogenic (Supplementary Table S3). One of these genes is *pncA*, the absence of which explains the pathogen’s resistance to PZA, as discussed previously. Another key gene in Del-X is *hspX*, which encodes an alpha-crystallin – a small heat-shock protein, which is essential for long-term viability during latent, asymptomatic infections [21]. The deletion of these 18 genes is thus expected to affect the pathogen’s physiology, in particular to impose a pressure against intra-phagosome existence, as further discussed in the discussion section.

### Closely related genomes

To find the origin of the *Mtb* strain that infected Mr. X, we utilized the NCBI Pathogen Detection platform [22] to identify closely related sequenced isolates. NCBI Pathogen Detection Project clusters related pathogen genome sequences in order to identify transmission networks and outbreaks. Based on genomic similarity, 13 *Mtb* isolates closely related to the strain from Mr. X were identified (Figure 3, Supplementary Table S4). These isolates were collected between 2007 and 2015 in Russia, Sweden, Moldova and Iran. The genome most closely related to the clone from Mr. X was collected in 2009 in Russia, differing from it by only four SNPs (Figure 3). None of these genomes contains Del-X, suggesting that the deletion event might have occurred in Mr. X immediately following his infection, or in another unknown host prior to his infection. Mr. X immigrated to Israel from Russia the same year in which he was diagnosed with active tuberculosis. Thus, it is reasonable to assume that he was infected in Russia, where a significant number of the isolates, which belong to the same cluster, were collected.

**Figure 3:**
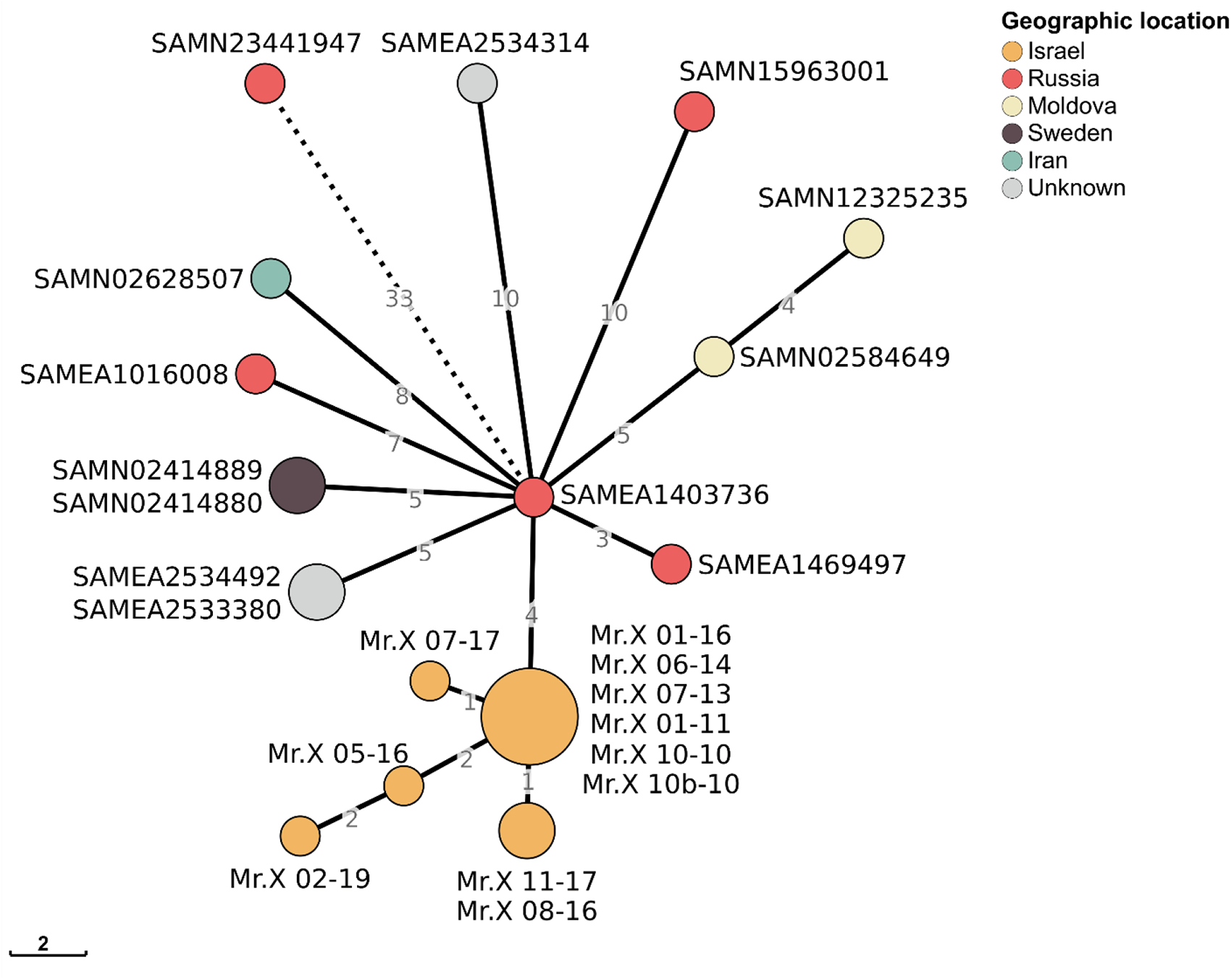
Minimum spanning tree of Mr. X isolates and other closely related genomes: Minimum spanning tree of 11 Mr. X’s samples and 13 closest samples from the Pathogen detection cluster PDS000010075.9. Numbers on branches represent the distance between nodes in SNPs, while nodes present a distance of zero SNPs. Nodes are colored according to their countries of origin. For visual purposes, the SAMN23441947-SAMEA1403736 branch has been shortened.

### Other genomes with deletions in Del-X region

In order to estimate the uniqueness of the Del-X deletion, we searched for deletions overlapping the Del-X region in all non-redundant, well-annotated, publicly available *Mtb* genomes. Among the 7028 RefSeq *Mtb* genomes found, 252 had deletions of various lengths overlapping Del-X (Supplementary File S5). Of these, 51 had a deletion longer than 1 kb, 33 had a deletion longer than 3 kb, and 22 had a deletion longer than 5 kb (Figure 4, Supplementary File S5). These genomes were isolated from various parts of the world, and belonged to different *Mtb* lineages, primarily the Beijing and the LAM lineages. It is important to notice that not all these 22 genomes represent unique deletion events. In fact, nine of them belong to three different clusters of nearly identical genomes. One such pair is particularly interesting: two clonal genomes with a 23.2 kb genomic deletion spanning almost the entire Del-X region, collected from a parent-child pair in Lima, Peru [23] (BioSamples SAMN10697670 and SAMN10697510, rows 4 and 5 in Figure 4). This observation suggests that despite its potential impact on dormancy, hypoxia response and intracellular survival, Del-X does not prevent host-to-host transmission. This pair of clonal isolates belong to a larger cluster in NCBI Pathogen Detection [22], comprising 223 isolates (cluster PDT000456105.1). Several of these 223 isolates have smaller deletions in Del-X region, such as a 194 bp deletion inside the Rv2028c gene, a deletion of 195 bp inside the *acg* gene (Rv2032), and 5 clonal isolates with a deletion spanning 7 genes inside Del-X, including *pncA*. However, the larger 23.2 kb genomic deletion is not likely to have emerged from any of these smaller deletions, as this region is intact in the genome, which is closest to the pair with this large deletion (BioSample SAMEA2683069). Similarly, BioSample SAMEA2682984, a strain that is clonal (0 SNPs in NCBI Pathogen Detection) to the cluster of strains with large deletions appearing in rows 12-16 of Figure 4, has no deletion in Del-X region.

**Figure 4:**
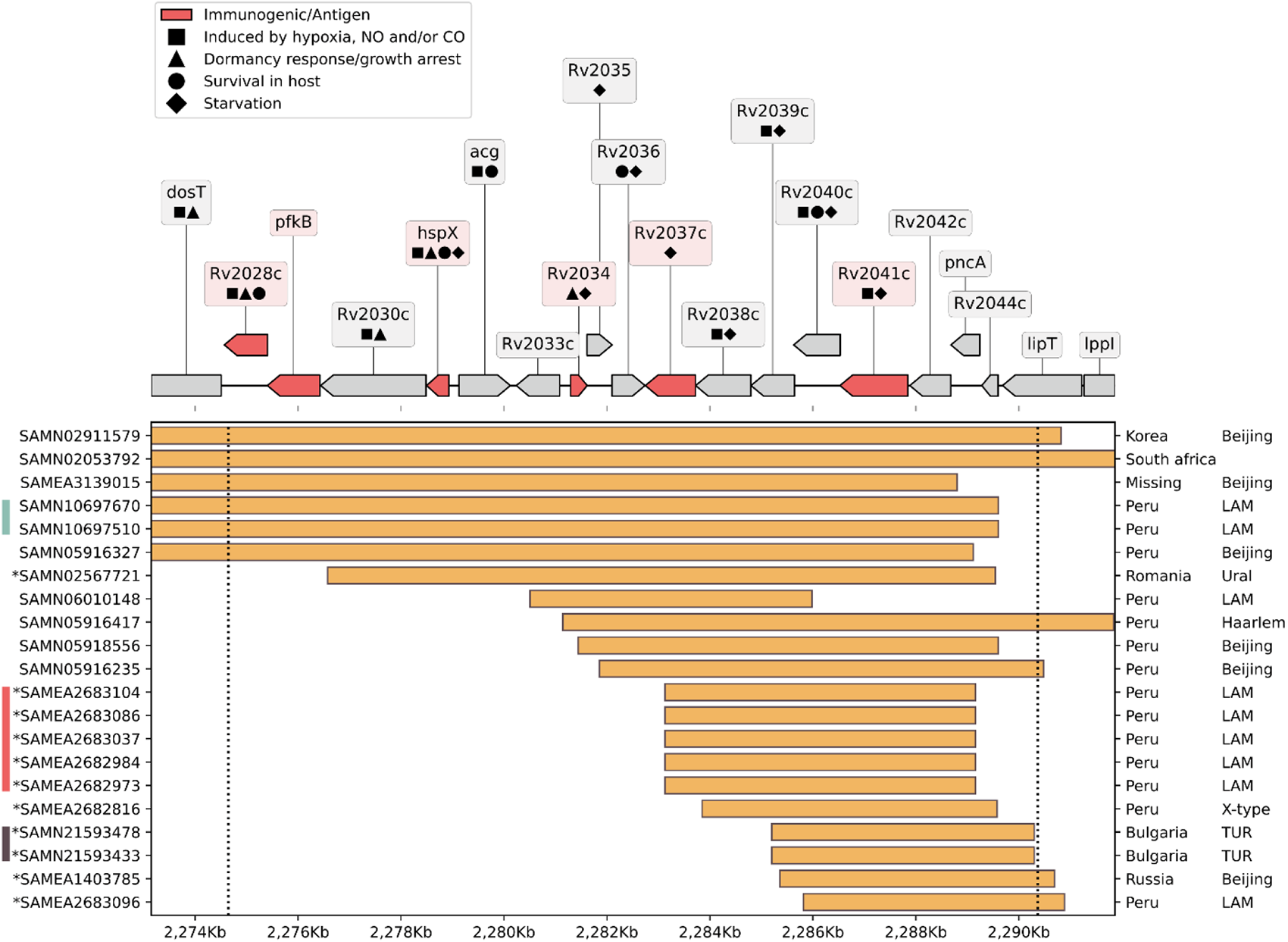
Deletions overlapping Del-X in other *Mtb* genomes: On the top: a genomic map of the Del-X region according to the H37Rv reference genome. Genes in red are immunogenic. Symbols represent gene functions of interest. On the bottom: Samples from the NCBI RefSeq containing a genomic deletion that overlaps the Del-X region. The deletions are shown as orange rectangles. Ticks on the right display country of origin and lineage. The left ticks indicate the BioSample accession number, and the asterisk denotes that the deletion was confirmed using the sequencing data from the SRA database. Bars to the left of BioSample IDs indicate identical deletions. Pathogen detection was used to find the number of SNP between samples with identical deletions. The turquoise bar shows the parent-child pair with one SNP difference. The samples marked with the red bar have zero distance between them. The brown bar shows two samples with 16 SNPs difference.

### *In vivo* evolution

All *Mtb* sequenced isolates from Mr. X were clonal. Nevertheless, due to the large deletion, the lack of adherence to therapy, exposure to multiple drugs and the comorbidities, the *Mtb* strain infecting Mr. X was subjected to unique selective pressures during the infection. We therefore utilized the pipeline developed by Gatt and Margalit [15], and identified 26 *Mtb* genes which underwent non-synonymous mutations during the eight and a half years of active tuberculosis. (Figure 5). Nine of these genes have been previously identified by Gatt and Margalit as ‘adaptive genes’, that tend to undergo changes at a relatively high frequency during in-host infection of *Mtb* [15], many of them belong to the PE-PGRS family. Among the 26 genes, seven underwent mutations predicted to lead to loss of function such as large deletions or frameshift mutations, and the remaining 19 genes underwent other non-synonymous mutations. The biological processes these genes are involved in are diverse, and include antibiotic resistance, virulence, fatty acid metabolism, central metabolism and more, reflecting the dynamic nature of bacterial adaptation to the *in vivo* environment.

**Figure 5:**
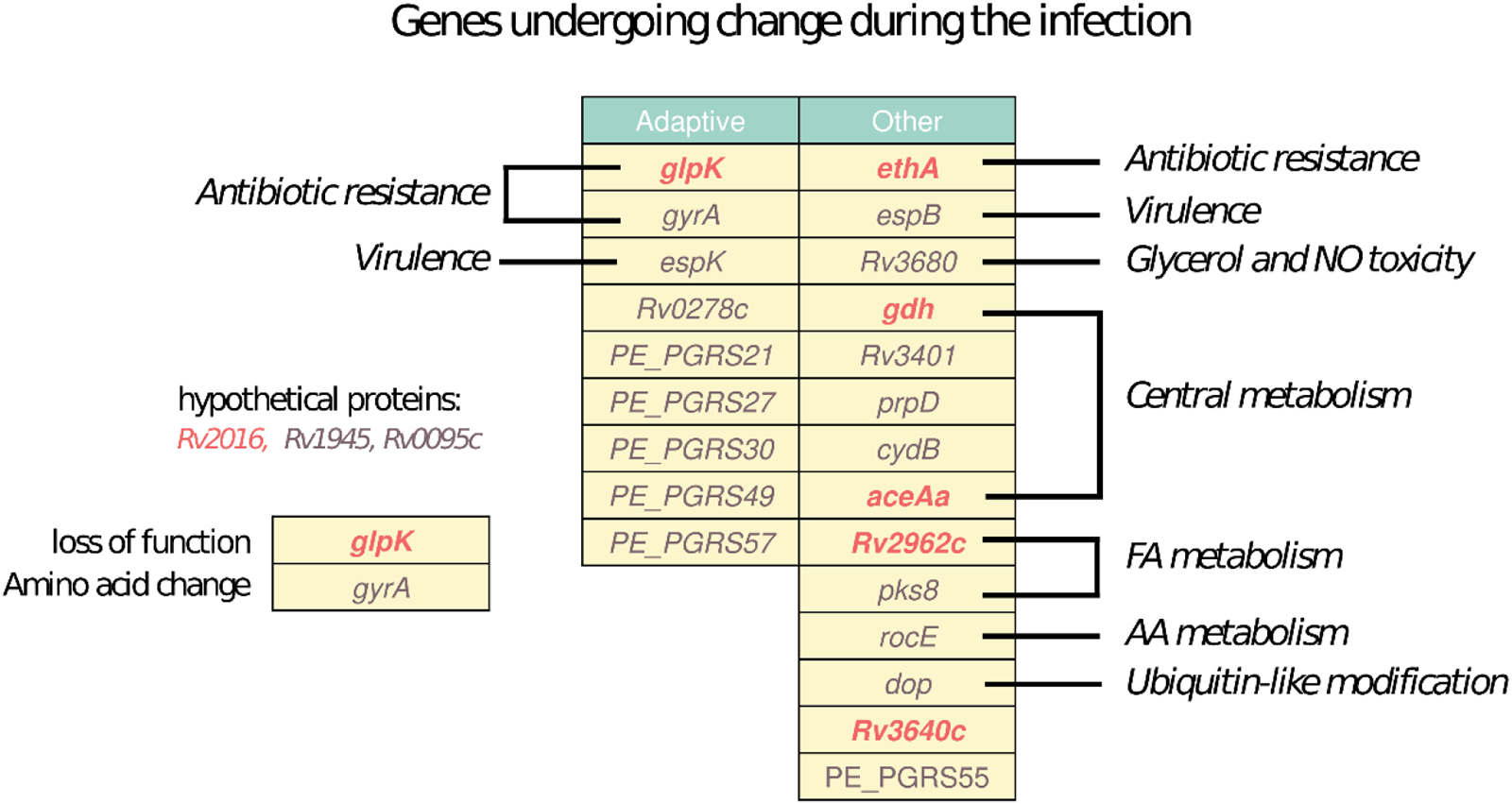
*In vivo* evolution: Table with genes determined to undergo non-synonymous mutations during the infection of Mr. X using the pipeline by Gatt and Margalit (2021). The left column includes genes determined as adaptive in *Mtb* by Gatt and Margalit (2021) and the right column includes additional genes. Functional categories the genes are associated with are denoted next to the table. Genes undergoing mutations in mechanisms predicted to lead to loss of function are marked in blue, and genes undergoing other non-synonymous mutations are marked in orange.

## Discussion

In this study, we present a case of exceptionally long survival in a man over 60 years of age who concurrently had progressive AIDS, active tuberculosis caused by MDR-TB and alcoholism. This remarkable extended survival is particularly notable, given the patient’s failure to complete his anti-TB treatment regimens, and his refusal of anti-HIV treatment as reflected by his very low CD4 cells counts and high viremic state. A previous study on survival rates in patients with drug resistance TB found a median survival of 5.9 years in MDR-TB and XDR-TB patients and 1.9 years in patients co-infected with HIV [8]. The advanced age, male gender and alcoholism of our patient makes this case even more exceptional [8]. The severe consequences and poor prognosis for patients co-infected with HIV/MDR-TB, particularly those that don’t take ART is well documented in the literature [24–27]. Furthermore, prior HIV infection, like in our case, significantly increase the chance of acquiring MDR-TB (1.42 times higher) [28].

Over the years, since his first TB diagnosis, Mr. X started treatment several times, but never completed it. He was treated with 13 different antibiotics in total. His treatment ultimately failed to cure his TB, presumably due to the combination of his co-morbidities, the pathogen’s multiple drug resistance and his low compliance (Figure 1, Supplementary Table S1). Initially the standard combination of RIF, INH, EMB and PZA was administered, but when DST results from the NMRL indicated that the strain was resistant to RIF, INH and PZA, these drugs were replaced with second line drugs. His treatment later included EMB again, despite most of his *Mtb* isolates exhibiting intermediate resistance to this anti-mycobacterial agent, and two isolate being fully resistant.

The isolation and genome sequencing of the *Mtb* isolates from this patient revealed an unusual genomic deletion, termed Del-X. This deletion spans from bases 2274648 to 2290386 in the H37Rv reference genome (RefSeq id NC_000962.3), causing the loss of 18 genes (Rv2028c to Rv2045c in the standard *Mtb* loci numbering). What effect could Del-X have on the bacterium’s physiology, and the nature of the disease? Many of the genes inside Del-X are involved in dormancy (Figure 2, Supplementary Table S3). The *hspX* gene (Rv2031c, also known as *acr*) encodes alpha-Crystallin-like heat shock protein (Hsp16.3), which plays a pivotal role in *Mtb* dormancy response [21,29,30]. DosR, a transcription factor that activates the transcription of *Mtb* dormancy regulon, was shown to induce the expression of *hspX* and *acg* [31]. In turn, Rv2028 and Rv2034 positively regulate the expression of the *dosR* gene [30,32]. Many of the genes within Del-X are induced by the conditions typical of macrophages phagosomes, such as hypoxia, starvation, low levels of nitric oxide, and acidic pH [30,33–39]. *hspX* actively contributes to slowing the growth of *Mtb* immediately following infection and is essential for its resistance to low oxygen stress [29,30]. Therefore, it is reasonable to speculate that the strain infecting Mr. X was unable to enter a dormant state. Studies have shown that a knockout strain of *Mtb* lacking *hspX* exhibits reduced growth in macrophages [21], and that depletion of HspX protein deteriorates *Mtb* tolerance to anaerobic conditions [30]. Rv2037c too was shown to enhance *in vivo* and *ex vivo* intracellular survival [37]. Thus, we suggest that this genomic deletion not only prevented the pathogen from entering a dormant state, but might have also abolished its ability to survive inside macrophages’ phagosomes, imposing an escape from the phagosomes into cells’ cytosol [40].

Several of the genes in Del-X region play an important role in the host immune response to *Mtb*. *hspX*, Rv2029c, Rv2034, Rv2041c, and possibly Rv2028c encode strong immunogenic antigens [41,42] (Figure 2, Supplementary Table S3). HspX in particular plays a major role in activation of dendritic cells and induces the production of TNF-α in lymphocytes [42,43]. *acg* is important for development of the typical tuberculosis granulomas [44]. In a study on MDR-TB patients, Rv2037c was shown to induce the production of antibodies and of pro-inflammatory cytokines [37]. Rv2041c increases the production of cytokines, which are important for macrophage maturation (M1 polarization) [45]. Therefore, Del-X is predicted to weaken the immune response against the pathogen, compared to the response against wild type *Mtb*. The patient’s immune system was severely compromised by his HIV infection, as evidenced by the low CD4 counts (about 80 per microliter), low CD4/CD8 ratio (10%), and high viral load (near 350,000 and 900,000 per milliliter) measured in 2017 and in 2018, respectively. The combined effect of impaired immune system and lack of *Mtb* determinants important for immunogenic recognition and activation, likely diminished the host ability to neutralize the infection, resulting in a constantly active TB.

On the other hand, Del-X may have severely affected the pathogen’s proliferation inside the host. Two of the deleted genes, Rv2028 and Rv2040c, have been previously identified in a comprehensive screen for genes that are important for in-host survival [46]. *acg* has also been shown to be an essential virulence factor, as its deletion attenuated the bacterial growth in acute and persistent mice infections [44]. The Rv2038c-Rv2041c operon encodes a sugar importer of the ABC transporter family, which is thought to play an important role in virulence or in-host survival [47], and its absence may limit the bacterial energy source. Furthermore, Del-X may impair lipid metabolism, due to the deletion of Rv2034 and Rv2037c [32,37], and nucleotide metabolism, due to the deletion of Rv2030c [30]. These deficiencies could further attenuate the growth of this strain. We hypothesize that Del-X genomic deletion resulted in an attenuated *Mtb* strain, unable to enter dormancy, yet with poor proliferation rate. The reduced virulence of this strain may have been a major factor contributing to the long survival of the patient. However, in contrast to this hypothesis, it is worth noting that some evidences have shown that deletion of *hspX* increases bacterial growth *in vivo* [29].

An obvious effect of Del-X was PZA resistance, due to the deletion of *pncA*. Treatment with PZA likely selected for the strain with this deletion, allowing it to take over the population, despite its defects. Indeed, although deletions in this genomic region are rare, almost all known *Mtb* genomes with a deletion of 5 kb or more in this region also exhibit a deletion of *pncA* (Figure 4, File S5). In line with this, the work of Maxime Godfroid *et al*, focusing on analysis of multidrug resistant MTB outbreak isolates, discovered that genomic insertions and deletions are significantly enriched in genes conferring antibiotic resistance [48].

The discovery of Del-X underscores the importance of detecting large genomic deletions in routine analysis of *Mtb* whole genome sequencing. Standard pipelines based on mapping to reference genomes and variant call may miss such deletions. Indeed, following this case, our laboratory has incorporated a step of identifying deletions based on uncovered genomic regions to its routine analysis.

We found a few *Mtb* genomes with genomic deletions in Del-X, from different parts of the world, mainly belonging to the Beijing or LAM lineages. As mentioned, the isolates from Mr. X also belong to the Beijing lineage that is known to be hyper-mutagenic, virulent, that rapidly acquires mutations for resistance to anti-tuberculosis drugs [49–52].

Interestingly, two of these genomes were isolated from a parent and their child, and are genetically clonal. Hence, one of these family members probably infected the other [23]. This finding suggests that the Del-X genomic deletion does not prevent transmission from one patient to another and may have already been present in the strain when Mr. X was infected. However, none of this strain’s known closely related isolates had a deletion in the Del-X region. Thus, we cannot exclude the possibility that the deletion arose within this patient (Figure 3, Supplementary Table S4).

The isolates obtained from Mr. X at different time points during his nearly nine years of infection belong to the same clone, yet there are several variations differing between them. These variations may originate from co-existing sub populations randomly sampled, or reflect changes that accumulated over time during the infection. Our phylogenetic analysis also attempts to separate these cases and include only mutations likely to have emerged during the infection. The later possibility is generally the case in *Mtb* infections [17]. Many of the genes mutated during the infection were previously identified as ‘hot spots’ for *in-vivo* evolution during TB infection (Figure 5) [15,16]. For example, the *gyrA* A90V mutation discussed above, which explains the emergence of FQ resistance in isolates obtained between August 2016 and November 2017, and was probably selected for by the treatment with MFX and LFX [53]. Furthermore, the fact that the patient did not receive continuous treatment is known to be a major risk factor for the appearance of resistance [5]. After the exposure to these drugs was removed, an isolate taken in February 2019 lacked this mutation, probably representing a sub-population that evaded FQs and became dominant once they were no longer used. Another interesting variation is the insertion of C in the homopolymeric tract of 7 cytosine in the *glpK* gene, relative to the TB reference H37Rv genome. This mutation is common to all isolates except for the one from August 2016 and the last isolate from February 2019. The *glpK* gene encodes a glycerol kinase involved in glycerol metabolism. This mutation is known to serve as a mechanism for drug tolerance and growth slow-down, which occurs and reverses at high frequency [54]. Notably, among the 26 genes mutated in-host there were six PE-PGRS genes, the functions of which are largely unknown (Figure 5).

The medical records of Mr. X indicate that he started TB treatment at least six different times, probably reflecting worsening of the symptoms (Figure 1). In all of these episodes, the patient stopped the treatment before completing it, at times by simply disappearing. This demonstrates the challenges faced by the health authorities in treating TB, which often involve social and personal difficulties. Treatment of low compliant and unmotivated patients is often unsuccessful and requires a tailored plan based on establishing a long-term trust relationship. Directly observed therapy (DOT) is an important part of such a plan [55].

Taken together, we hypothesize that the unusual prolonged TB illness of Mr. X was a result of a unique intricate interplay between the host and the pathogen. The potentially low immunogenic signature of the pathogen together with the severe immune deficiency of the host prevented the elimination of the pathogen. The Del-X genomic deletion prevented latency, resulting in constant active TB. At the same time, Del-X probably resulted in an attenuated pathogen, allowing the long-term survival of the patient.

The importance of this report stems from the unusual long survival of Mr. X, considering the circumstances, as well as the uniqueness of the genomic deletion described here. This case raises important questions about the influence of *Mtb* genetic factors on disease progression and the potential implications for prognosis and treatment. It is interesting to find out whether in additional cases of patients who survive active tuberculosis for many years without treatment, mutated *Mtb* with genomic mutations in important genes are involved. Further investigations may elucidate the precise mechanisms by which the Del-X deletion affects the pathogenicity and virulence of *Mtb*, as well as the host immune response to the infection. In addition, this special isolate, can be used as a model bacterium for a better understanding of host-pathogen fundamental interactions, such as *Mtb* escape from the phagosome to the cytosol, dormancy and more.

## Materials and methods

### Ethics statement

This study was approved by the National Helsinki Committee for Medical Experiments on Humans, of the Israeli Ministry of Health (# MOH-081-2021).

### Data classification and identification of *Mycobacterium tuberculosis complex* (MTBC)

All cultures processed in Israel are sent to the National Mycobacterium Reference Laboratory (NMRL), in Tel Aviv, Israel. At the NMRL, smears from samples and TB cultures are stained using Ziehl-Neelsen staining and cultured on Lowenstein-Jensen (LJ) media using standard methods [56], and in addition cultured on BACTECe MGITe 960 (BD, Sparks, MD, USA) system. The species are identified using conventional biochemical methods [57], [58] and a commercially available strip DNA probe test (Hain Lifesciences, Nehren, Germany).

### Drug susceptibility testing (DST) for MTBC strains

DST for all the drugs was done using resistance ratio method [59]. Pyrazinamide (PZA) was tested using Mark’s stepped pH method [60].

Drugs threshold levels used to determine resistance are described in Supplementary Table S6. Importantly, for the purpose of the article, the categories Borderline and RR4 were redefined as Intermediate.

### Whole genome sequencing and Bioinformatics

Genomic DNA from inactivated bacterial cultures was extracted according to the manufacturer’s protocol using Maxwell RSC cultured cells DNA kit and Maxwell® 16 System (Promega) [61]. DNA Paired-end libraries were prepared using the Illumina Nextera XT DNA Library Preparation Kit according to Illumina protocols. For sequencing, we utilized the Illumina MiSeq platform using a MiSeq Reagent Kit v2 (500-cycles) (catalogue MS-102-2003) or a MiSeq Reagent Kit v3 (600-cycle), (catalogue MS-102-3003).

Raw reads fastq files were quality analyzed by FastQC [62], Kraken2 taxonomic classification [63] was used to identify mixed cultures. MTBseq, a comprehensive pipeline for whole genome sequence analysis of MTBC isolates, was used as the main tool for lineage annotation, identification of genomic variations and clusters analysis [64]. Lineages were identified as part of the MTBseq pipeline, based on the SNP scheme developed by Coll et al [65]. Genomic variations were compared to the WHO 2021 Catalogue of drug resistance associated mutations in MTBC and used to identify resistant strains to antibiotics [20].

### Searching for deletions among assemblies from the NCBI

A search for deletions was carried out on 7298 *Mtb* (taxid1773) assemblies available in the NCBI RefSeq database [66] (accessed March 15 2023). After excluding replaced and anomalous assemblies, 7028 valid assemblies remained. Assembly metadata were retrieved using the Entrez Direct [https://www.ncbi.nlm.nih.gov/books/NBK179288/], and sequences were downloaded using the ncbi-genome-download v0.3.1 tool [https://github.com/kblin/ncbi-genome-download]. All 7028 sequences were then aligned to the reference sequence of *Mtb* H37Rv (NC_000962.3) using the NUCmer program from the MUMmer v4.0.0rc1 package [67] with the default options. Assembly containing uncovered regions of any length that overlap with H37Rv region 2274648 – 2290368 was considered to have a deletion (Supp_File_S5).

### Deletions verification with sequence read archive (SRA) data

Samples with assembly-predicted deletions longer than 1kb were additionally evaluated using sequencing data from the SRA database [68]. 32 samples out of 51 with total deletion sizes of more than 1 kb have sequencing data in the SRA. The GRIDSS v2.13.2 software for the detection of genomic rearrangements [69,70] was used to examine these samples. Additionally, Samtools v1.16.1 [71] was used to calculate per position read depth for all samples. The deletion is considered to be correctly verified if: 1) The deletion predicted by GRIDSS overlaps with the deletion predicted from the assembly by more than 99% of the length and vice versa; 2) More than 90% of the bases in the given region have a read depth of less than 5% of the mean genome depth. Assembly GCF_000679915.1 had 4 SRA runs, 3 of them passed, therefore we considered this assembly as verified. After applying these filters seven samples were discarded. Samples that lack sequencing data in the SRA database were used in the further analysis without verification (Supp_File_S5).

### Long reads

For Oxford Nanopore sequencing, DNA from Mr. X ten isolate (except 02-19) were converted into a sequencing library using Rapid Barcoding Kit 96 (SQK-RBK110.96) chemistry according to the manufacturer’s protocol. Sequencing was performed on a MinION sequencer with an R9.4.1 (FLO-MIN106D) flow cell and basecalling was carried out by Guppy version 6.0.1. Sequences of all 11 isolates were used to create a single assembly.

### MST construction

13 samples from the Pathogen detection cluster PDS000010075.9 and 11 Mr. X’s isolates were used to construct the minimum spanning tree. There are 3 samples from Pathogen detection (BioSample accession numbers SAMN23441947, SAMN15963001, and SAMN12325235) for which there are no sequencing data in the SRA database [68]. Assembled sequences of these samples were artificially converted to paired-end reads using a custom python script. The forward and reverse read lengths were both set to 150bp with a minimal coverage of 50. Prefetch and fasterq-dump from the SRA Toolkit v3.0.1 [https://github.com/ncbi/sra-tools/] were used to obtain FASTQ files from the remaining 10 samples. BWA-MEM v0.7.17-r1188 [72] was used to map the reads to the *Mtb* H37Rv (NC_000962.3) reference sequence. Viewing, sorting, and duplication removal in BAM files were done using Samtools v1.16.1 [71]. FreeBayes v1.3.6 [73] was used to call SNPs with the -p 1 -q 3 parameters. SNPs were filtered using the following criteria: Site quality >= 30, variant read depth >= 5, distance to nearest SNP >= 3 bp, >= 5% read mapping up-or downstream, >= 5% read mapping to least-covered strand, variant allele frequency >= 90%. In addition, SNPs in genes associated with antibiotic resistance as well as those located in PE/PPE were excluded in the subsequent analysis [74]. Sites with more than 5% of missing variants were filtered out as the final stage of filtering. SNP filtering was carried out using VCFtools v 0.1.16 [75], bcftools v1.16 [71], vcffilter from vcflib v 1.0.3 [76], and bedtools v2.30.0 [77]. The GrapeTree v 1.5.0 [78] with MSTree method was used to create the minimum spanning tree.

### Lineage assignment

Genome assemblies were mapped to the reference *Mtb* H37Rv genome (RefSeq id NC_000962.3), and variants were detected as previously described [79]. These variants were compared to a SNP barcode of *Mtb* lineages to assign a lineage [65].

### Determination of within-host evolution

Detection of mutations was performed as in Gatt and Margalit [15]. Briefly, assemblies were constructed for all paired-end libraries using kSNP v.3.1 [80,81], and phylogenetic structure was determined using the TRACE algorithm with high confidence progenitor-progeny isolate pairs ascertained from the different isolates of Mr. X. For each progenitor-progeny pair, *breseq* v.0.32 [82,83] was then used to determine differential variations from the reference genome *Mtb* H37Rv (RefSeq id NC_000962.3) appearing in the progeny isolate and not in the progenitor isolate. SnpEff v.4.1 [84] was used to predict the effect of the different mutations, with high impact mutations predicted to lead to loss of function.

## Data availability

Genomic sequence data was deposited to NCBI database under BioProject PRJNA834625.

## Author contribution

MR, IN, AM: conceived the study concept, conceived and designed the bioinformatic analysis, analyzed data. MR and IN wrote the manuscript. YL: analyzed phenotypic data and helped write the manuscript. YEG, HM: perform the *in-vivo* bioinformatics analysis and critically reviewed the manuscript. OM, DAZ: performed the ONT sequencing, bioinformatics analysis and critically read the manuscript. IK, GZV: conducted the whole genome sequencing and critically read the manuscript. ZD, HKS: critical reading of the manuscript. MS, LT: collected and summarize medical records of Mr.X and critically read of the manuscript. ER: project supervision and critical reading of the manuscript. All authors read and approved the final manuscript.

## Bibliography

1. Organization, World Health. Global tuberculosis report 2022. *Geneva*: *World Health Organization*; 2022. *Licence: CC BY-NC-SA 3.0 IGO*. 2022: Geneva

2. WHO operational handbook on tuberculosis. *Module 4: treatment – drug-resistant tuberculosis treatment, 2022 update. Geneva: World Health Organization*; 2022. Licence: CC BY-NC-SA 3.0 IGO.

3. Sotgiu, G.; Sulis, G.; Matteelli, A. Tuberculosis-a World Health Organization Perspective. Microbiol Spectr 2017. 5(1) DOI: 10.1128/microbiolspec.TNMI7-0036-2016.

4. Linh, N. N.; Viney, K.; Gegia, M.; Falzon, D.; Glaziou, P.; Floyd, K.; Timimi, H.; Ismail, N.; Zignol, M.; Kasaeva, T.; Mirzayev, F. World Health Organization treatment outcome definitions for tuberculosis: 2021 update. Eur Respir J 2021. 58(2) DOI: 10.1183/13993003.00804-2021.

5. Gillespie, S. H. Evolution of drug resistance in Mycobacterium tuberculosis: clinical and molecular perspective. Antimicrob Agents Chemother 2002. 46(2), 267–74 DOI: 10.1128/AAC.46.2.267-274.2002.

6. Bell, L. C. K.; Noursadeghi, M. Pathogenesis of HIV-1 and Mycobacterium tuberculosis co-infection. Nat Rev Microbiol 2018. 16(2), 80–90 DOI: 10.1038/nrmicro.2017.128.

7. Nordholm, A. C.; Andersen, A. B.; Wejse, C.; Norman, A.; Ekstrom, C. T.; Andersen, P. H.; Lillebaek, T.; Koch, A. Mortality, risk factors, and causes of death among people with tuberculosis in Denmark, 1990-2018. Int J Infect Dis 2023. 130, 76–82 DOI: 10.1016/j.ijid.2023.02.024.

8. Balabanova, Y.; Ignatyeva, O.; Fiebig, L.; Riekstina, V.; Danilovits, M.; Jaama, K.; Davidaviciene, E.; Radiulyte, B.; Popa, C. M.; Nikolayevskyy, V.; Drobniewski, F. Survival of patients with multidrug-resistant TB in Eastern Europe: what makes a difference? Thorax 2016. 71(9), 854–61 DOI: 10.1136/thoraxjnl-2015-207638.

9. Wigger, G. W.; Bouton, T. C.; Jacobson, K. R.; Auld, S. C.; Yeligar, S. M.; Staitieh, B. S. The Impact of Alcohol Use Disorder on Tuberculosis: A Review of the Epidemiology and Potential Immunologic Mechanisms. Front Immunol 2022. 13, 864817 DOI: 10.3389/fimmu.2022.864817.

10. Singh, P. R.; Vijjamarri, A. K.; Sarkar, D. Metabolic Switching of Mycobacterium tuberculosis during Hypoxia Is Controlled by the Virulence Regulator PhoP. J Bacteriol 2020. 202(7) DOI: 10.1128/JB.00705-19.

11. Leistikow, R. L.; Morton, R. A.; Bartek, I. L.; Frimpong, I.; Wagner, K.; Voskuil, M. I. The Mycobacterium tuberculosis DosR regulon assists in metabolic homeostasis and enables rapid recovery from nonrespiring dormancy. J Bacteriol 2010. 192(6), 1662–70 DOI: 10.1128/JB.00926-09.

12. Peddireddy, V.; Doddam, S. N.; Ahmed, N. Mycobacterial Dormancy Systems and Host Responses in Tuberculosis. Front Immunol 2017. 8, 84 DOI: 10.3389/fimmu.2017.00084.

13. Delogu, G.; Sali, M.; Fadda, G. The biology of mycobacterium tuberculosis infection. Mediterr J Hematol Infect Dis 2013. 5(1), e2013070 DOI: 10.4084/MJHID.2013.070.

14. Verma, A.; Ghoshal, A.; Dwivedi, V. P.; Bhaskar, A. Tuberculosis: The success tale of less explored dormant Mycobacterium tuberculosis. Front Cell Infect Microbiol 2022. 12, 1079569 DOI: 10.3389/fcimb.2022.1079569.

15. Gatt, Y. E.; Margalit, H. Common Adaptive Strategies Underlie Within-Host Evolution of Bacterial Pathogens. Mol Biol Evol 2021. 38(3), 1101–1121 DOI: 10.1093/molbev/msaa278.

16. Ley, S. D.; de Vos, M.; Van Rie, A.; Warren, R. M. Deciphering Within-Host Microevolution of Mycobacterium tuberculosis through Whole-Genome Sequencing: the Phenotypic Impact and Way Forward. Microbiol Mol Biol Rev 2019. 83(2) DOI: 10.1128/MMBR.00062-18.

17. Gagneux, S. Ecology and evolution of Mycobacterium tuberculosis. Nat Rev Microbiol 2018. 16(4), 202–213 DOI: 10.1038/nrmicro.2018.8.

18. Dos Vultos, T.; Mestre, O.; Rauzier, J.; Golec, M.; Rastogi, N.; Rasolofo, V.; Tonjum, T.; Sola, C.; Matic, I.; Gicquel, B. Evolution and diversity of clonal bacteria: the paradigm of Mycobacterium tuberculosis. PLoS One 2008. 3(2), e1538 DOI: 10.1371/journal.pone.0001538.

19. Baena, A.; Cabarcas, F.; Ocampo, J. C.; Barrera, L. F.; Alzate, J. F. Large genomic deletions delineate Mycobacterium tuberculosis L4 sublineages in South American countries. PLoS One 2023. 18(5), e0285417 DOI: 10.1371/journal.pone.0285417.

20. Catalogue of mutations in Mycobacterium tuberculosis complex and their association with drug resistance. Geneva: World Health Organization; 2021. *Licence: CC BY-NC-SA 3.0 IGO*.

21. Yuan, Y.; Crane, D. D.; Simpson, R. M.; Zhu, Y. Q.; Hickey, M. J.; Sherman, D. R.; Barry, C. E., 3rd The 16-kDa alpha-crystallin (Acr) protein of Mycobacterium tuberculosis is required for growth in macrophages. Proc Natl Acad Sci U S A 1998. 95(16), 9578–83 DOI: 10.1073/pnas.95.16.9578.

22. The NCBI Pathogen Detection Project [Internet]. Bethesda (MD): National Library of Medicine (US), National Center for Biotechnology Information. 2016 May. Available from: https://www.ncbi.nlm.nih.gov/pathogens/.

23. Dixit, A.; Freschi, L.; Vargas, R.; Calderon, R.; Sacchettini, J.; Drobniewski, F.; Galea, J. T.; Contreras, C.; Yataco, R.; Zhang, Z.; Lecca, L.; Kolokotronis, S. O.; Mathema, B.; Farhat, M. R. Whole genome sequencing identifies bacterial factors affecting transmission of multidrug-resistant tuberculosis in a high-prevalence setting. Sci Rep 2019. 9(1), 5602 DOI: 10.1038/s41598-019-41967-8.

24. Bayowa, J. R.; Kalyango, J. N.; Baluku, J. B.; Katuramu, R.; Ssendikwanawa, E.; Zalwango, J. F.; Akunzirwe, R.; Nanyonga, S. M.; Amutuhaire, J. S.; Muganga, R. K.; Cherop, A. Mortality rate and associated factors among patients co-infected with drug resistant tuberculosis/HIV at Mulago National Referral Hospital, Uganda, a retrospective cohort study. PLOS Glob Public Health 2023. 3(7), e0001020 DOI: 10.1371/journal.pgph.0001020.

25. Chem, E. D.; Van Hout, M. C.; Hope, V. Treatment outcomes and antiretroviral uptake in multidrug-resistant tuberculosis and HIV co-infected patients in Sub Saharan Africa: a systematic review and meta-analysis. BMC Infect Dis 2019. 19(1), 723 DOI: 10.1186/s12879-019-4317-4.

26. Isaakidis, P.; Casas, E. C.; Das, M.; Tseretopoulou, X.; Ntzani, E. E.; Ford, N. Treatment outcomes for HIV and MDR-TB co-infected adults and children: systematic review and meta-analysis. Int J Tuberc Lung Dis 2015. 19(8), 969–78 DOI: 10.5588/ijtld.15.0123.

27. Lelisho, M. E.; Wotale, T. W.; Tareke, S. A.; Alemu, B. D.; Hassen, S. S.; Yemane, D. M.; Korsa, B. B.; Bedaso, N. G. Survival rate and predictors of mortality among TB/HIV co-infected adult patients: retrospective cohort study. Sci Rep 2022. 12(1), 18360 DOI: 10.1038/s41598-022-23316-4.

28. Sultana, Z. Z.; Hoque, F. U.; Beyene, J.; Akhlak-Ul-Islam, M.; Khan, M. H. R.; Ahmed, S.; Hawlader, D. H.; Hossain, A. HIV infection and multidrug resistant tuberculosis: a systematic review and meta-analysis. BMC Infect Dis 2021. 21(1), 51 DOI: 10.1186/s12879-020-05749-2.

29. Hu, Y.; Movahedzadeh, F.; Stoker, N. G.; Coates, A. R. Deletion of the Mycobacterium tuberculosis alpha-crystallin-like hspX gene causes increased bacterial growth in vivo. Infect Immun 2006. 74(2), 861–8 DOI: 10.1128/IAI.74.2.861-868.2006.

30. Mushtaq, K.; Sheikh, J. A.; Amir, M.; Khan, N.; Singh, B.; Agrewala, J. N. Rv2031c of Mycobacterium tuberculosis: a master regulator of Rv2028-Rv2031 (HspX) operon. Front Microbiol 2015. 6, 351 DOI: 10.3389/fmicb.2015.00351.

31. Park, H. D.; Guinn, K. M.; Harrell, M. I.; Liao, R.; Voskuil, M. I.; Tompa, M.; Schoolnik, G. K.; Sherman, D. R. Rv3133c/dosR is a transcription factor that mediates the hypoxic response of Mycobacterium tuberculosis. Mol Microbiol 2003. 48(3), 833–43 DOI: 10.1046/j.1365-2958.2003.03474.x.

32. Gao, C. H.; Yang, M.; He, Z. G. Characterization of a novel ArsR-like regulator encoded by Rv2034 in Mycobacterium tuberculosis. PLoS One 2012. 7(4), e36255 DOI: 10.1371/journal.pone.0036255.

33. Betts, J. C.; Lukey, P. T.; Robb, L. C.; McAdam, R. A.; Duncan, K. Evaluation of a nutrient starvation model of Mycobacterium tuberculosis persistence by gene and protein expression profiling. Mol Microbiol 2002. 43(3), 717–31 DOI: 10.1046/j.1365-2958.2002.02779.x.

34. Cassio Barreto de Oliveira, M.; Balan, A. The ATP-Binding Cassette (ABC) Transport Systems in Mycobacterium tuberculosis: Structure, Function, and Possible Targets for Therapeutics. Biology (Basel) 2020. 9(12) DOI: 10.3390/biology9120443.

35. Garbe, T. R.; Hibler, N. S.; Deretic, V. Response to reactive nitrogen intermediates in Mycobacterium tuberculosis: induction of the 16-kilodalton alpha-crystallin homolog by exposure to nitric oxide donors. Infect Immun 1999. 67(1), 460–5 DOI: 10.1128/IAI.67.1.460-465.1999.

36. Kim, S. Y.; Lee, B. S.; Shin, S. J.; Kim, H. J.; Park, J. K. Differentially expressed genes in Mycobacterium tuberculosis H37Rv under mild acidic and hypoxic conditions. J Med Microbiol 2008. 57(Pt 12), 1473–1480 DOI: 10.1099/jmm.0.2008/001545-0.

37. Kumari, B.; Saini, V.; Kaur, J.; Kaur, J. Rv2037c, a stress induced conserved hypothetical protein of Mycobacterium tuberculosis, is a phospholipase: Role in cell wall modulation and intracellular survival. Int J Biol Macromol 2020. 153, 817–835 DOI: 10.1016/j.ijbiomac.2020.03.037.

38. Saxena, A.; Srivastava, V.; Srivastava, R.; Srivastava, B. S. Identification of genes of Mycobacterium tuberculosis upregulated during anaerobic persistence by fluorescence and kanamycin resistance selection. Tuberculosis (Edinb) 2008. 88(6), 518–25 DOI: 10.1016/j.tube.2008.01.003.

39. Snasel, J.; Machova, I.; Solinova, V.; Kasicka, V.; Krecmerova, M.; Pichova, I. Phosphofructokinases A and B from Mycobacterium tuberculosis Display Different Catalytic Properties and Allosteric Regulation. Int J Mol Sci 2021. 22(3) DOI: 10.3390/ijms22031483.

40. Jamwal, S. V.; Mehrotra, P.; Singh, A.; Siddiqui, Z.; Basu, A.; Rao, K. V. Mycobacterial escape from macrophage phagosomes to the cytoplasm represents an alternate adaptation mechanism. Sci Rep 2016. 6, 23089 DOI: 10.1038/srep23089.

41. Legesse, M.; Ameni, G.; Medhin, G.; Mamo, G.; Franken, K. L.; Ottenhoff, T. H.; Bjune, G.; Abebe, F. IgA response to ESAT-6/CFP-10 and Rv2031 antigens varies in patients with culture-confirmed pulmonary tuberculosis, healthy Mycobacterium tuberculosis-infected and non-infected individuals in a tuberculosis endemic setting, Ethiopia. Scand J Immunol 2013. 78(3), 266–74 DOI: 10.1111/sji.12080.

42. Meier, N. R.; Battegay, M.; Ottenhoff, T. H. M.; Furrer, H.; Nemeth, J.; Ritz, N. HIV-Infected Patients Developing Tuberculosis Disease Show Early Changes in the Immune Response to Novel Mycobacterium tuberculosis Antigens. Front Immunol 2021. 12, 620622 DOI: 10.3389/fimmu.2021.620622.

43. Amir, M.; Aqdas, M.; Nadeem, S.; Siddiqui, K. F.; Khan, N.; Sheikh, J. A.; Agrewala, J. N. Diametric Role of the Latency-Associated Protein Acr1 of Mycobacterium tuberculosis in Modulating the Functionality of Pre- and Post-maturational Stages of Dendritic Cells. Front Immunol 2017. 8, 624 DOI: 10.3389/fimmu.2017.00624.

44. Hu, Y.; Coates, A. R. Mycobacterium tuberculosis acg gene is required for growth and virulence in vivo. PLoS One 2011. 6(6), e20958 DOI: 10.1371/journal.pone.0020958.

45. Su-Young Kim, A-Rum Shin, Byung-Soo Lee, Hwa-Jung Kim Bo Young Jeon, Sang-Nae Cho, Jeong-Kyu Park, Sung Jae Shin Characterization of Immune Responses to Mycobacterium tuberculosis Rv2041c Protein. Journal of Bacteriology and Virology 2009. 39(3), 183–193 DOI: DOI: 10.4167/jbv.2009.39.3.183.

46. Sassetti, C. M.; Boyd, D. H.; Rubin, E. J. Genes required for mycobacterial growth defined by high density mutagenesis. Mol Microbiol 2003. 48(1), 77–84 DOI: 10.1046/j.1365-2958.2003.03425.x.

47. De la Torre, L. I.; Vergara Meza, J. G.; Cabarca, S.; Costa-Martins, A. G.; Balan, A. Comparison of carbohydrate ABC importers from Mycobacterium tuberculosis. BMC Genomics 2021. 22(1), 841 DOI: 10.1186/s12864-021-07972-w.

48. Godfroid, M.; Dagan, T.; Merker, M.; Kohl, T. A.; Diel, R.; Maurer, F. P.; Niemann, S.; Kupczok, A. Insertion and deletion evolution reflects antibiotics selection pressure in a Mycobacterium tuberculosis outbreak. PLoS Pathog 2020. 16(9), e1008357 DOI: 10.1371/journal.ppat.1008357.

49. Ford, C. B.; Shah, R. R.; Maeda, M. K.; Gagneux, S.; Murray, M. B.; Cohen, T.; Johnston, J. C.; Gardy, J.; Lipsitch, M.; Fortune, S. M. Mycobacterium tuberculosis mutation rate estimates from different lineages predict substantial differences in the emergence of drug-resistant tuberculosis. Nat Genet 2013. 45(7), 784–90 DOI: 10.1038/ng.2656.

50. Hakamata, M.; Takihara, H.; Iwamoto, T.; Tamaru, A.; Hashimoto, A.; Tanaka, T.; Kaboso, S. A.; Gebretsadik, G.; Ilinov, A.; Yokoyama, A.; Ozeki, Y.; Nishiyama, A.; Tateishi, Y.; Moro, H.; Kikuchi, T.; Okuda, S.; Matsumoto, S. Higher genome mutation rates of Beijing lineage of Mycobacterium tuberculosis during human infection. Sci Rep 2020. 10(1), 17997 DOI: 10.1038/s41598-020-y.

51. Liu, Q.; Wang, D.; Martinez, L.; Lu, P.; Zhu, L.; Lu, W.; Wang, J. Mycobacterium tuberculosis Beijing genotype strains and unfavourable treatment outcomes: a systematic review and meta-analysis. Clin Microbiol Infect 2020. 26(2), 180–188 DOI: 10.1016/j.cmi.2019.07.016.

52. Ribeiro, S. C.; Gomes, L. L.; Amaral, E. P.; Andrade, M. R.; Almeida, F. M.; Rezende, A. L.; Lanes, V. R.; Carvalho, E. C.; Suffys, P. N.; Mokrousov, I.; Lasunskaia, E. B. Mycobacterium tuberculosis strains of the modern sublineage of the Beijing family are more likely to display increased virulence than strains of the ancient sublineage. J Clin Microbiol 2014. 52(7), 2615–24 DOI: 10.1128/JCM.00498-14.

53. Singh, R.; Dwivedi, S. P.; Gaharwar, U. S.; Meena, R.; Rajamani, P.; Prasad, T. Recent updates on drug resistance in Mycobacterium tuberculosis. J Appl Microbiol 2020. 128(6), 1547–1567 DOI: 10.1111/jam.14478.

54. Safi, H.; Gopal, P.; Lingaraju, S.; Ma, S.; Levine, C.; Dartois, V.; Yee, M.; Li, L.; Blanc, L.; Ho Liang, H. P.; Husain, S.; Hoque, M.; Soteropoulos, P.; Rustad, T.; Sherman, D. R.; Dick, T.; Alland, D. Phase variation in Mycobacterium tuberculosis glpK produces transiently heritable drug tolerance. Proc Natl Acad Sci U S A 2019. 116(39), 19665–19674 DOI: 10.1073/pnas.1907631116.

55. World Health Organization. Communicable Diseases Cluster. (1999). *What is DOTS? : a guide to understanding the WHO-recommended TB control strategy known as DOTS. World Health Organization.* https://apps.who.int/iris/handle/10665/65979.

56. Kent, P. T.; Kubica, G. P., Public Health Mycobacteriology: A Guide for the Level III Laboratory. 1985 pages.

57. Marks, J. A system for the examination of tubercle bacilli and other mycobacteria. Tubercle 1976. 57(3), 207–25 DOI: 10.1016/0041-3879(76)90030-1.

58. Collins, C. H.; Grange, J. M.; Yates, M. D., Organization and Practice in Tuberculosis Bacteriology. 1985 pages.

59. *European Centre for Disease Prevention and Control. Handbook on tuberculosis laboratory diagnostic methods in the European Union – Updated 2018.* *Stockholm*: ECDC; 2018.

60. Marks, J. A ‘Stepped Ph’ Technique for the Estimation of Pyrazinamide Sensitivity. Tubercle 1964. 45, 47–50 DOI: 10.1016/s0041-3879(64)80087-8.

61. Cabibbe, A. M.; Walker, T. M.; Niemann, S.; Cirillo, D. M. Whole genome sequencing of Mycobacterium tuberculosis. Eur Respir J 2018. 52(5) DOI: 10.1183/13993003.01163-2018.

62. Andrews, S. FASTQC. A quality control tool for high throughput sequence data. 2010.

63. Wood, D. E.; Lu, J.; Langmead, B. Improved metagenomic analysis with Kraken 2. Genome Biol 2019. 20(1), 257 DOI: 10.1186/s13059-019-1891-0.

64. Kohl, T. A.; Utpatel, C.; Schleusener, V.; De Filippo, M. R.; Beckert, P.; Cirillo, D. M.; Niemann, S. MTBseq: a comprehensive pipeline for whole genome sequence analysis of Mycobacterium tuberculosis complex isolates. PeerJ 2018. 6, e5895 DOI: 10.7717/peerj.5895.

65. Coll, F.; McNerney, R.; Guerra-Assuncao, J. A.; Glynn, J. R.; Perdigao, J.; Viveiros, M.; Portugal, I.; Pain, A.; Martin, N.; Clark, T. G. A robust SNP barcode for typing Mycobacterium tuberculosis complex strains. Nat Commun 2014. 5, 4812 DOI: 10.1038/ncomms5812.

66. O’Leary, N. A.; Wright, M. W.; Brister, J. R.; Ciufo, S.; Haddad, D.; McVeigh, R.; Rajput, B.; Robbertse, B.; Smith-White, B.; Ako-Adjei, D.; Astashyn, A.; Badretdin, A.; Bao, Y.; Blinkova, O.; Brover, V.; Chetvernin, V.; Choi, J.; Cox, E.; Ermolaeva, O.; Farrell, C. M.; Goldfarb, T.; Gupta, T.; Haft, D.; Hatcher, E.; Hlavina, W.; Joardar, V. S.; Kodali, V. K.; Li, W.; Maglott, D.; Masterson, P.; McGarvey, K. M.; Murphy, M. R.; O’Neill, K.; Pujar, S.; Rangwala, S. H.; Rausch, D.; Riddick, L. D.; Schoch, C.; Shkeda, A.; Storz, S. S.; Sun, H.; Thibaud-Nissen, F.; Tolstoy, I.; Tully, R. E.; Vatsan, A. R.; Wallin, C.; Webb, D.; Wu, W.; Landrum, M. J.; Kimchi, A.; Tatusova, T.; DiCuccio, M.; Kitts, P.; Murphy, T. D.; Pruitt, K. D. Reference sequence (RefSeq) database at NCBI: current status, taxonomic expansion, and functional annotation. Nucleic Acids Res 2016. 44(D1), D733–45 DOI: 10.1093/nar/gkv1189.

67. Marcais, G.; Delcher, A. L.; Phillippy, A. M.; Coston, R.; Salzberg, S. L.; Zimin, A. MUMmer4: A fast and versatile genome alignment system. PLoS Comput Biol 2018. 14(1), e1005944 DOI: 10.1371/journal.pcbi.1005944.

68. Leinonen, R.; Sugawara, H.; Shumway, M.; International Nucleotide Sequence Database, Collaboration The sequence read archive. Nucleic Acids Res 2011. 39(Database issue), D19-21 DOI: 10.1093/nar/gkq1019.

69. Cameron, D. L.; Baber, J.; Shale, C.; Valle-Inclan, J. E.; Besselink, N.; van Hoeck, A.; Janssen, R.; Cuppen, E.; Priestley, P.; Papenfuss, A. T. GRIDSS2: comprehensive characterisation of somatic structural variation using single breakend variants and structural variant phasing. Genome Biol 2021. 22(1), 202 DOI: 10.1186/s13059-021-02423-x.

70. Cameron, D. L.; Schroder, J.; Penington, J. S.; Do, H.; Molania, R.; Dobrovic, A.; Speed, T. P.; Papenfuss, A. T. GRIDSS: sensitive and specific genomic rearrangement detection using positional de Bruijn graph assembly. Genome Res 2017. 27(12), 2050–2060 DOI: 10.1101/gr.222109.117.

71. Danecek, P.; Bonfield, J. K.; Liddle, J.; Marshall, J.; Ohan, V.; Pollard, M. O.; Whitwham, A.; Keane, T.; McCarthy, S. A.; Davies, R. M.; Li, H. Twelve years of SAMtools and BCFtools. Gigascience 2021. 10(2) DOI: 10.1093/gigascience/giab008.

72. Li H. (2013) Aligning sequence reads, clone sequences and assembly contigs with BWA-MEM. arXiv:1303.3997v1 [q-bio.GN].

73. Garrison, E.; Marth, G. T. Haplotype-based variant detection from short-read sequencing. 2012, DOI: 10.48550/arXiv.1207.3907.

74. Kohl, T. A.; Diel, R.; Harmsen, D.; Rothganger, J.; Walter, K. M.; Merker, M.; Weniger, T.; Niemann, S. Whole-genome-based Mycobacterium tuberculosis surveillance: a standardized, portable, and expandable approach. J Clin Microbiol 2014. 52(7), 2479–86 DOI: 10.1128/JCM.00567-14.

75. Danecek, P.; Auton, A.; Abecasis, G.; Albers, C. A.; Banks, E.; DePristo, M. A.; Handsaker, R. E.; Lunter, G.; Marth, G. T.; Sherry, S. T.; McVean, G.; Durbin, R.; Genomes Project Analysis, Group The variant call format and VCFtools. Bioinformatics 2011. 27(15), 2156–8 DOI: 10.1093/bioinformatics/btr330.

76. Garrison, E.; Kronenberg, Z. N.; Dawson, E. T.; Pedersen, B. S.; Prins, P. A spectrum of free software tools for processing the VCF variant call format: vcflib, bio-vcf, cyvcf2, hts-nim and slivar. PLoS Comput Biol 2022. 18(5), e1009123 DOI: 10.1371/journal.pcbi.1009123.

77. Quinlan, A. R.; Hall, I. M. BEDTools: a flexible suite of utilities for comparing genomic features. Bioinformatics 2010. 26(6), 841–2 DOI: 10.1093/bioinformatics/btq033.

78. Zhou, Z.; Alikhan, N. F.; Sergeant, M. J.; Luhmann, N.; Vaz, C.; Francisco, A. P.; Carrico, J. A.; Achtman, M. GrapeTree: visualization of core genomic relationships among 100,000 bacterial pathogens. Genome Res 2018. 28(9), 1395–1404 DOI: 10.1101/gr.232397.117.

79. Rubinstein, M.; Grossman, R.; Nissan, I.; Schwaber, M. J.; Carmeli, Y.; Kaidar-Shwartz, H.; Dveyrin, Z.; Rorman, E. Mycobacterium intracellulare subsp. chimaera from Cardio Surgery Heating-Cooling Units and from Clinical Samples in Israel Are Genetically Unrelated. Pathogens 2021. 10(11) DOI: 10.3390/pathogens10111392.

80. Gardner, S. N.; Hall, B. G. When whole-genome alignments just won’t work: kSNP v2 software for alignment-free SNP discovery and phylogenetics of hundreds of microbial genomes. PLoS One 2013. 8(12), e81760 DOI: 10.1371/journal.pone.0081760.

81. Gardner, S. N.; Slezak, T.; Hall, B. G. kSNP3.0: SNP detection and phylogenetic analysis of genomes without genome alignment or reference genome. Bioinformatics 2015. 31(17), 2877–8 DOI: 10.1093/bioinformatics/btv271.

82. Barrick, J. E.; Colburn, G.; Deatherage, D. E.; Traverse, C. C.; Strand, M. D.; Borges, J. J.; Knoester, D. B.; Reba, A.; Meyer, A. G. Identifying structural variation in haploid microbial genomes from short-read resequencing data using breseq. BMC Genomics 2014. 15(1), 1039 DOI: 10.1186/1471-2164-15-1039.

83. Deatherage, D. E.; Barrick, J. E. Identification of mutations in laboratory-evolved microbes from next-generation sequencing data using breseq. Methods Mol Biol 2014. 1151, 165–88 DOI: 10.1007/978-1-4939-0554-6_12.

84. Cingolani, P.; Platts, A.; Wang le, L.; Coon, M.; Nguyen, T.; Wang, L.; Land, S. J.; Lu, X.; Ruden, D. M. A program for annotating and predicting the effects of single nucleotide polymorphisms, SnpEff: SNPs in the genome of Drosophila melanogaster strain w1118; iso-2; iso-3. Fly (Austin) 2012. 6(2), 80–92 DOI: 10.4161/fly.19695.

85. Li, H. Minimap2: pairwise alignment for nucleotide sequences. Bioinformatics 2018. 34(18), 3094–3100 DOI: 10.1093/bioinformatics/bty191.

